# Frameshift variants in *TBX2* underlie autosomal-dominant hearing loss with incomplete penetrance of nystagmus

**DOI:** 10.1101/2024.07.18.24310488

**Authors:** Wan Hua, Yanfei Wang, Xiang Li, Lanchen Wang, Wenyu Xiong, Meilin Chen, Fengxiao Bu, Libo Liu, Mingjun Zhong, Yu Lu, Zhiyong Liu, Jing Cheng, Huijun Yuan

## Abstract

**Purpose:** The transcription factor TBX2 plays a critical role in inner hair cells development in mice. Yet, the link between *TBX2* malfunction and human hearing-related disorders remains unexplored.

**Methods:** Linkage analysis combined with whole genome sequencing was applied to identify the causative gene in two autosomal dominant Chinese families characterized by late-onset progressive sensorineural hearing loss and incomplete penetrance of horizontal oscillatory nystagmus. Functional evaluation of *TBX2* variants was performed through protein expression, localization, and transcriptional activity analysis *in vitro*, phenotypic analysis and mechanism study in knockout mice model *in vivo*.

**Results:** Multipoint parametric linkage analysis of Family 1 revealed a maximum LOD score of 3.01 on chromosome 17q23.2. Whole genome sequencing identified distinct *TBX2* variants, c.977delA (p.Asp326Alafs*42) and c.987delC (p.Ala330Argfs*38) in each family, co-segregating with hearing loss. These variants resulted in premature termination and the generation of a new peptide segment, reducing transcriptional activity. Further, heterozygous *Tbx2* knockout mice exhibited late-onset progressive hearing loss, along with ectopic expression of Prestin in IHCs and a gradual decrease in expression from P7 to P42.

**Conclusion:** Our findings indicate that heterozygous *TBX2* frameshift variants are the genetic cause of late-onset progressive hearing loss and incomplete penetrance of nystagmus. The heterozygous *Tbx2* knockout mouse model mirrored the human hearing loss phenotype, further validating TBX2’s role in auditory function. These insights enhance our understanding of *TBX2* in the auditory system, providing valuable information for molecular diagnostics and genetic counseling in related hearing disorders.

## Introduction

Hearing loss (HL), influenced by both environmental and genetic factors, affects over 1.5 billion people globally, with projections indicating a rise to 2.5 billion by 2050 in WHO report (1). Genetic factors account for 60-80% of congenital hearing loss, with 40-50% linked to known HL genes, while a substantial proportion of patients remain undiagnosed (2–4). The diagnosis rate of late-onset HL, particularly syndromic HL with additional phenotypes, is lower due to genetic and phenotypic heterogeneity as well as environmental influences (5–7). Despite these challenges, the discovery of new pathogenic genes is crucial for clinical diagnosis and understanding the mechanisms of the auditory system (1, 8).

The auditory epithelium comprises two types of sound receptor cells, Inner Hair Cells (IHCs) and Outer Hair Cells (OHCs), crucial for auditory function (9, 10). Recently, Tbx2 has been identified as a key regulator of IHCs development, controlling inner versus outer hair cell differentiation, and shaping the IHCs and inner supporting cells (ISCs) of sensory epithelium (11–13). The expression of Tbx2 promotes the transdifferentiation of cochlear supporting cells and OHCs into IHCs, significantly advancing cochlear regeneration research (14, 15). Tbx2 is widely expressed in mice cochlea, with particularly high expression in IHCs and other epithelial cell types lining the cochlear scala media (11). TBX2, encoding the T-box transcription factor, contains a conserved T-box domain that binds to DNA sequence-specific regions, functioning as a transcriptional activator and repressor (16, 17). It is a critical regulator of cardiac, skeletal, and neural development (18–20). Several large deletions encompassing *TBX2* have been reported associated with human developmental disorders, skeletal malformations, and cardiac defects (21–23). Although *TBX2* variants are linked to various disorders, such as conotruncal heart defects (MIM:217095) (24), vertebral anomalies and variable endocrine and T-cell dysfunction (MIM:618223) (25), as well as osteochondrodysplasia (26, 27), the association between TBX2 dysfunction and human hearing loss remains unproven, rendering the role of TBX2 in the human cochlea uncertain.

In this study, we aimed to identify the genetic cause of a new disorder characterized by progressive, post-lingual hearing loss combined with incomplete penetrance of nystagmus. We discovered two heterozygous frameshift variants in *TBX2* linked to this condition in two independent Chinese families. Robust genetic evidence was provided to confirm the pathogenicity of *TBX2* variant in this new disease in humans. Furthermore, based on the results from *in vitro* and *in vivo* studies in heterozygous *Tbx2* knockout mice, it is suggested that the mechanism by which *TBX2* causes hearing loss might be related to haploinsufficiency.

## Materials and Methods

### Subjects and clinical evaluations

The Chinese Deafness Genetics Consortium (CDGC) project, launched in September 2013, which encompassing 22,125 patients from 20,665 affected families. Family 1 and 2 were recruited from the Fujian and Hebei Provinces of mainland China, respectively. This study received approval from the Ethics Committee of West China Hospital (No.2020-606). All participants and/or their legal guardians provided written informed consent in accordance with the guidelines set by the ethics committees and institutional review boards of the respective institutions.

All participants underwent various audiological tests, such as pure-tone audiometry (PTA), acoustic impedance, auditory brainstem response (ABR), distortion product of otoacoustic emission (DPOAE), and otolaryngological examination. PTA was employed to determine hearing threshold levels (dB HL) for air and bone conduction at multiple frequencies (250, 500, 1000, 2000, 4000, and 8000 Hz). The severity of hearing loss was categorized based on the average thresholds across specific frequencies (500, 1000, 2000, and 4000 Hz), with classification criteria across the range of normal (<20 dB), mild (20-40 dB), moderate (41-70 dB), severe (71-95 dB), and profound (>95 dB) deafness. Selected individuals from Family 1 underwent additional investigations, including computed tomography (CT) scan of the temporal bone, vestibular testing, visual functional assessments (e.g., signal field analysis, visual evoked potential (VEP)), blood and urine analyses, color Doppler ultrasonography of renal arteries, and echocardiography. Binaural mean air conduction thresholds (dB HL) were recorded for individuals aged 5 to 70 years, with hearing levels varying between 20 and 120 dB based on age and frequency.

### Screening of known genes underlying hearing loss

Each participant in the CDGC initially underwent screening using a SNP scan assay (Shanghai Genesky Biotech, Shanghai, China), which targeted 96 single nucleotide variants (SNVs), 19 insertions/deletions (indels), and three copy number variation (CNV) loci within GJB2, SLC26A4, and MT-RNR1. Subsequently, individuals without a genetic diagnosis underwent sequencing of the exons and ±50 flanking bases of 157 hearing loss-related genes using the CDGC-Hearing Loss panel based on the Agilent SureSelect Target Enrichment assay. Massively parallel sequencing (MPS) was conducted on Illumina sequencers, and DNA variants were called following the Genome Analysis Toolkit software (GATK) best practices workflow (28).

### Whole-genome linkage analysis

Genomic DNA was extracted from the peripheral blood samples obtained from 33 available members of Family 1 using a MagPure Tissue & Blood DNA LQ Kit (Magen, China). Genotyping and whole-genome linkage analysis were carried out using an Illumina Infinium Asian Screening Array MD-24 (Illumina, USA; Guoke Biotechnology, China), encompassing 690,000 high-density single-nucleotide polymorphism (SNP) markers. Multipoint parametric linkage analysis was conducted in Merlin version 1.1.2, assuming a dominant inheritance model for the disease phenotype. SNPs with a call rate below 95% and those displaying Mendelian inconsistencies were excluded from the analysis. The logarithm of odds (LOD) was computed with a mutant allele prevalence frequency of 0.0001 and penetrance values of 0.01, 0.90, and 0.90 for wild type homozygotes, mutant heterozygotes, and mutant homozygotes, respectively.

### Whole genome sequencing (WGS) and pathogenicity analysis

Whole genome sequencing was conducted on 24 members of Family 1 (including 13 individuals with hearing loss) and four members of Family 2, using the DNBSEQ-T7 platform (BGI, Shenzhen, China) with paired-end 150 base reads. Reads were mapped along the human reference genome (GRCh38/hg38) with the Burrows-Wheeler Aligner (version 0.7.15), variants were called using the Genomic Analysis Tool Kit, and Variant Effect Predictor was utilized for variant annotation.

A stepwise filtering strategy was employed to identify causative variants in patients. Variants meeting criteria of reading depth≥6, genotype quality≥20, and minor allele frequency (MAF) thresholds of 0.005 in public databases were selected for further analysis. Subsequently, variants within coding and non-coding regions predicted to affect protein function. As not all patients with hearing loss have accompanying nystagmus, a separate analysis was conducted on Family 1 with nystagmus as an independent phenotype using the aforementioned filtering strategy based on WGS analysis under both autosomal recessive and autosomal dominant pattern, respectively. Previously reported variants in known hearing loss and nystagmus genes were identified through public databases such as DVD (Deafness Variation Database), ClinVar, OMIM, HPO, and literature. A systematic literature review was conducted on all mutant genes to ascertain their potential connection with hearing loss and nystagmus, considering the patient’s phenotype and the effect of candidate genes on inner ear function based on *in vivo* or *in vitro* experiments. To mitigate misinterpretation due to variant types or sequencing quality, the Integrative Genomics Viewer (IGV) v.2.8.11 was utilized to manually review diagnostic variants.

### Sanger sequencing

DNA samples from all available family members (34 from Family 1; 10 from Family 2) underwent Sanger sequencing to assess whether potential variants in identified genes co-segregated with the disease phenotype. The DNA sequence containing TBX2 candidate variants was amplified by PCR using the following primers: Forward 5’-ACAACAACCCGTTTGCCAAG-3’; Reverse 5’-CCTTCCTGGCCTTTCAGGTT-3’. The PCR products were sequenced bidirectionally using the ABI 3500xL Dx Genetic Analyzer (Applied Biosystems, Thermo Fisher Scientific). Variant genotypes were confirmed through sequence analysis using Chromas.

### Cell culture, Western blot, immunofluorescence staining, and dual luciferase-based transcription reporter assays

Human embryonic kidney 293 cells (HEK293T) and Hela cells were purchased from American Type Culture Collection (ATCC, Manassas, Virginia, United States) and cultured in Dulbecco’s modified Eagle medium DMEM (SH30022.01B, HyClone, Logan, UT, United States) with 10% fetal bovine serum (26140079, Gibco, United States). A plasmid expressing WT TBX2, N-terminally attached to Flag, was synthesized from Sangon Biotech (Shanghai, China). The *TBX2* c.977delA, p.Asp326Alafs*42 and c.987delC, p.Ala330Argfs*38 were generated by site-directed mutagenesis KLD reactions (M0554, NEB). The cells were harvested for TBX2 expression analysis by Western blot after transfecting for 48 h. The antibodies used included anti-Flag (F1804, Sigma-Aldrich) and GAPDH (D190090, Sangon Biotech, Shanghai, China). GAPDH was used as the protein loading control. For immunofluorescence staining, Hela cells were transfected with TBX2 plasmids with 24 h. The antibodies included anti-Flag (F1804, Sigma-Aldrich), Alexa Fluor 555 Phalloidin (A34055, Thermo Fisher Scientific), and goat anti-mouse Alexa Fluor 647 secondary antibody (A48289, Thermo Fisher Scientific). Fluorescence images were captured by confocal microscopy (STELLARIS 5, Leica Microsystems). For dual luciferase transcription reporter assay, we constructed a reporter plasmid based on pGL3-promoter (HG-VQP0124, Promega,) that containing four palindromic T-box binding site sequences (AATTTCACACCTAGGTGTGAAATT), pRL-TK (E2231, Promega) was used as the internal reference reporter plasmid. Transcriptional activity assay was performed according to the manufacturer’s instructions of Dual-Luciferase® Reporter Assay System (Promega, E1910). Each assay was performed in technical duplicates and repeated at least three times.

### Generation and genotyping of *Tbx2* heterozygous knockout mice

*Tbx2^+/−^* mice in the C57BL/6 background were created using CRISPR/Cas9, as described in a previous study (12). Two small-guide RNAs (sgRNA): 5’-TGACCCGCCGTAAGGGCCTG-3’ (sgRNA-1) and 5’-AGGCTCCGAGGCGCCGACGT-3’ (sgRNA-2) were combined with Cas9 mRNA and injected into the cytoplasm of C57BL/6J zygotes. To confirm the sequence of the mutant allele, Founder 0 (F0) mice were identified through genotyping PCR (Tbx2-F, 5’-CTCCCTCTGAAGTGCATGGA-3’, Tbx2-R1, 5’-CAGGGAGAAGGTGTCGGAAG-3’; Tbx2-R2, 5’-GAAAGTCGCCAGCAAGTTGA-3’). Subsequently, accurately sequenced F0 mice were bred with C57BL/6 wild type mice to produce F1 mice, which underwent genotyping PCR and were utilized for the functional investigations in this study.

### Auditory brainstem response (ABR) assessment

Auditory brainstem response (ABR) analysis was carried out at frequencies of 4 kHz, 5.6 kHz, 8 kHz, 11.3 kHz, 16 kHz, 22.6 kHz, and 32 kHz in anesthetized mice aged P70 to P150. Mice were deeply anesthetized via intraperitoneal injection of 480 mg/kg chloral hydrate and placed on a heating pad to maintain a body temperature of 37 ℃. Three subdermal needle electrodes were positioned: one at the vertex (active electrode), another at the right mastoid region (reference electrode), and the third at the left shoulder (ground electrode). ABR measurements were controlled using BioSigRZ software. The simulation involved tone-burst short pure tone signals lasting 3 ms, with a 1 ms rise and fall time, delivered at a frequency of 20 per second. Recordings were conducted for 15 ms and averaged 400 times. Sound intensity ranged from 90 dB SPL to 0 dB SPL, decrementing by 5 dB.

### Immunofluorescence assay and calculation of inner hair cells

The dissection and immunofluorescent staining of inner ear sample from P7, P14, and P42 were followed the previously reported protocol (12). Primary antibodies used included goat polyclonal anti-Prestin (sc-22692, Santa Cruz) and rabbit polyclonal anti-vGlut3 (135203, Synaptic Systems). Immunofluorescence images were acquired using Nikon C2 (Nikon, Japan), TiE-A1 plus (Nikon, Japan), or NiE-A1 plus (Nikon, Japan) confocal microscopes.

A detailed scan of three segments of the cochlear duct was performed by using a 10x lens with confocal microscope. The Image J software was employed to draw lines between IHCs and OHCs to calculate the total length of these three segments. Following that, each cochlea was evenly divided into three equal-length portions, these portions were categorized as the basal, middle, or apical turns base on respective locations. We conducted a scan of the entire cochlea using a confocal microscope with a 60x lens to quantify the proportion of IHC expressing Prestin (Prestin+ IHCs), and total IHCs that expressing vGlut3 (vGlut3+ IHCs). The percentage of Prestin+ IHCs was calculated by normalizing numbers of all Prestin+ IHCs to total IHCs.

### Statistical analysis

Data were analyzed utilizing GraphPad Prism 8.0. Experiments were analyzed using an unpaired Student’s *t*-test and one-way ANOVA. A significance level of *p* < 0.05 was deemed statistically significant.

## Results

### Clinical features of patients

Family 1 spans five generations and comprises 102 members with hearing loss in autosomal-dominant inheritance pattern. The onset of hearing loss ranged from 4 to 40 years, with most affected members experiencing onset between 7 and 15 years (Table 1). Audiograms taken at a 10-year interval of three patients indicated a slowly progressive auditory decline (Figure 1A). Hearing loss was observed in 21 family members, characterized by symmetrical, mild to severe, progressive loss, initially affecting high frequencies and subsequently extending to all frequencies (Figure 1B). The audiograms of affected members displayed a sloping contour. Audiologic evaluations demonstrated normal immittance testing and bone conduction values equivalent to air conduction measurements, suggesting sensorineural hearing loss. Severe tinnitus and vertigo were self-reported by some members after hearing loss onset (Table 1). Notably, nine family members exhibited spontaneous nystagmus (Video S1), with eight also experiencing hearing loss (Table 1). Nystagmus onset occurred between 0-5 years old, preceding hearing loss onset. Visual pathway conduction defects were observed in three patients with both hearing loss and nystagmus, as indicated by visual evoked potential (VEP) clinical test.

**Figure 1.**
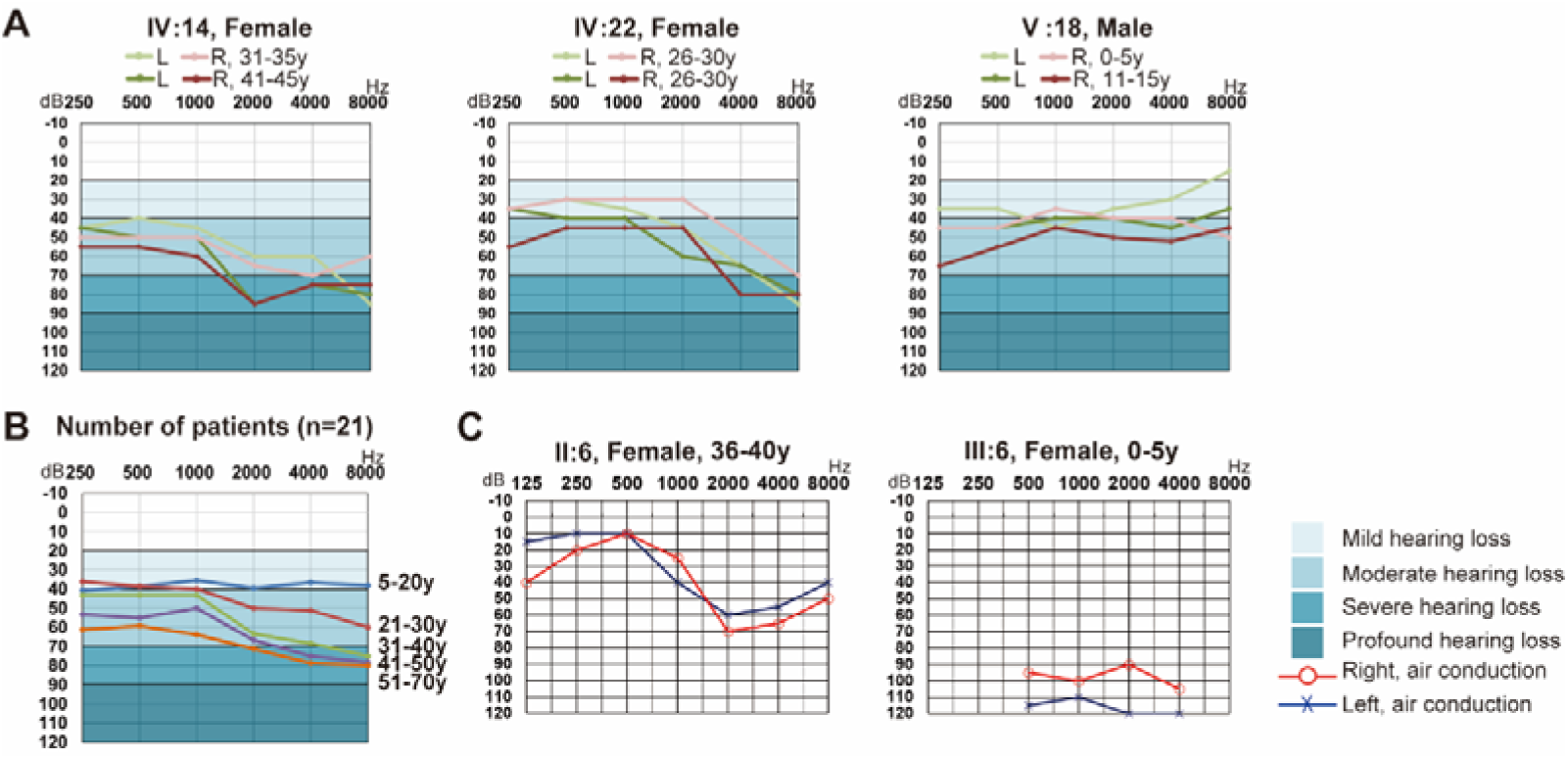
Audiograms of Family 1 and 2. A. Pure tone audiometry (PTA) of 3 patients from Family 1 at an interval of ten years; L: left ear, R: right ear. B. Age-related typical audiograms (ARTA) of Family 1. C: PTA of 2 patients from Family 2.

**Table 1.** Summary of clinical features for the affected individuals of Family 1 and Family 2.

| Family | Patient | Gender | PTA* |  | Audiogram shape | Level of hearing impairment | Tinnitus | Vertigo | Nystagmus |
| --- | --- | --- | --- | --- | --- | --- | --- | --- | --- |
|  |  |  | Left ear | Right ear |  |  |  |  |  |
| Family 1 | III:2 | Female | 69 | 88 | Sloping | Moderate | Yes | Yes | No |
|  | III:7 | Male | 80 | 100 | Sloping | Moderate | NA | NA | No |
|  | III:12 | Female | 68 | 84 | Sloping | Moderate | Yes | No | No |
|  | IV:11 | Male | 43 | 56 | Flat | Moderate | No | No | No |
|  | IV:14 | Female | 51 | 59 | Sloping | Moderate | Yes | No | No |
|  |  | Female | 65 | 69 | Sloping | Moderate | Yes | No | No |
|  | IV:16 | Female | 48 | 54 | Sloping | Moderate | No | No | No |
|  | IV:19 | Female | 61 | 70 | Sloping | Moderate | No | Yes | No |
|  | IV:22 | Female | 44 | 35 | Sloping | Moderate | No | Yes | Yes |
|  |  | Female | 51 | 54 | Sloping | Moderate | No | Yes | Yes |
|  | IV:23 | Female | 44 | 40 | Flat | Moderate | Yes | No | Yes |
|  | IV:27 | Male | 50 | 55 | Flat | Moderate | No | No | Yes |
|  | IV:31 | Male | 70 | 55 | Sloping | Moderate | Yes | No | Yes |
|  | IV:36 | Male | 50 | 53 | Sloping | Moderate | Yes | No | Yes |
|  | V:11 | Female | 41 | 35 | Flat | Mild | No | No | No |
|  | V:14 | Male | 43 | 50 | Flat | Moderate | No | No | No |
|  | V:15 | Male | 36 | 49 | Sloping | Mild | No | No | No |
|  | V:18 | Male | 36 | 40 | Flat | Mild | NA | NA | No |
|  |  | Male | 43 | 51 | Flat | Moderate | NA | NA | No |
|  | V:19 | Female | 32 | 33 | Flat | Mild | No | No | No |
|  | V:20 | Male | 39 | 40 | Sloping | Mild | No | No | Yes |
|  | V:22 | Female | 37 | 25 | Flat | Mild | NA | NA | Yes |
|  | V:23 | Female | 48 | 47 | Flat | Moderate | NA | NA | No |
|  | V:24 | Male | 28 | 36 | Flat | Mild | NA | NA | No |
| Family 2 | II:6 | Female | 30 | 31 | Sloping | Mild | No | No | Yes |
|  | III:6 | Female | 116 | 98 | Sloping | Profound | No | No | No |

\*The degree of hearing loss was defined according to pure-tone averages (PTAs), which
were based on the frequencies 500, 1000, 2000, and 4000 Hz.
NA: not available.

Family 2 includes 14 members over three generations, with four individuals exhibiting hearing loss. The proband was diagnosed with severe hearing loss between 0-5 years old, and has used hearing aids since diagnosis (Figure 1C). Pure-tone audiometry evaluation of patient II:6 presented progressive bilateral sensorineural symmetric and mild hearing loss, with late onset between 26-30 years old, consistent with Family 1’s audiological characteristics. The potential impact of pneumonia medication during pregnancy on the proband’s hearing loss cannot be ruled out, as viral infections during pregnancy could lead to hearing loss. Nystagmus was observed in the patient II:6 during childhood between 0-5 years old, preceding hearing loss onset, similar to Family 1.

No abnormalities were detected in blood tests, urine test, color Doppler ultrasonography of renal arteries, or echocardiography (Family 1). CT scans of the temporal bones showed no anomalies in the inner ears or vestibular systems of any examined patients (Family 1, Family 2). Comprehensive physical examinations, including head and neck palpation and interviews with affected individuals or their parents, did not reveal additional symptoms affecting multiple organs. Body weight and height, motor and neuronal development, as well as behavior and cognition, were within normal range.

### Identification of *TBX2* variants

Target genome sequencing for 157 HL related genes excluded the possibility of known HL genes in both families. To identify the locus of hearing loss, multipoint parametric linkage analysis of 33 individuals (21 with hearing loss) from Family 1 achieved a maximal logarithm of the odds (LOD) score of 3.01 for chr17q23.2: 41,511,650-66,337,413 (GRCh38/hg38) (Figure 2A). WGS was conducted for 13 affected and 10 unaffected members from Family 1 (Table S1). Filtering variants by quality control (Read depth≥6, genotype quality≥20), minor allele frequency (MAF) thresholds of 0.005 in public database, functional consequences (e.g., pLoF, missense, inframe indel), and autosomal dominant inheritance pattern (Figure 2B, Table S1) identified NM_005994.4, c.977delA, p.Asp326Alafs*42 in *TBX2* as co-segregating with the hearing loss phenotype, confirmed via Sanger sequencing, and within the critical linkage region (Figure 2C).

**Figure 2.**
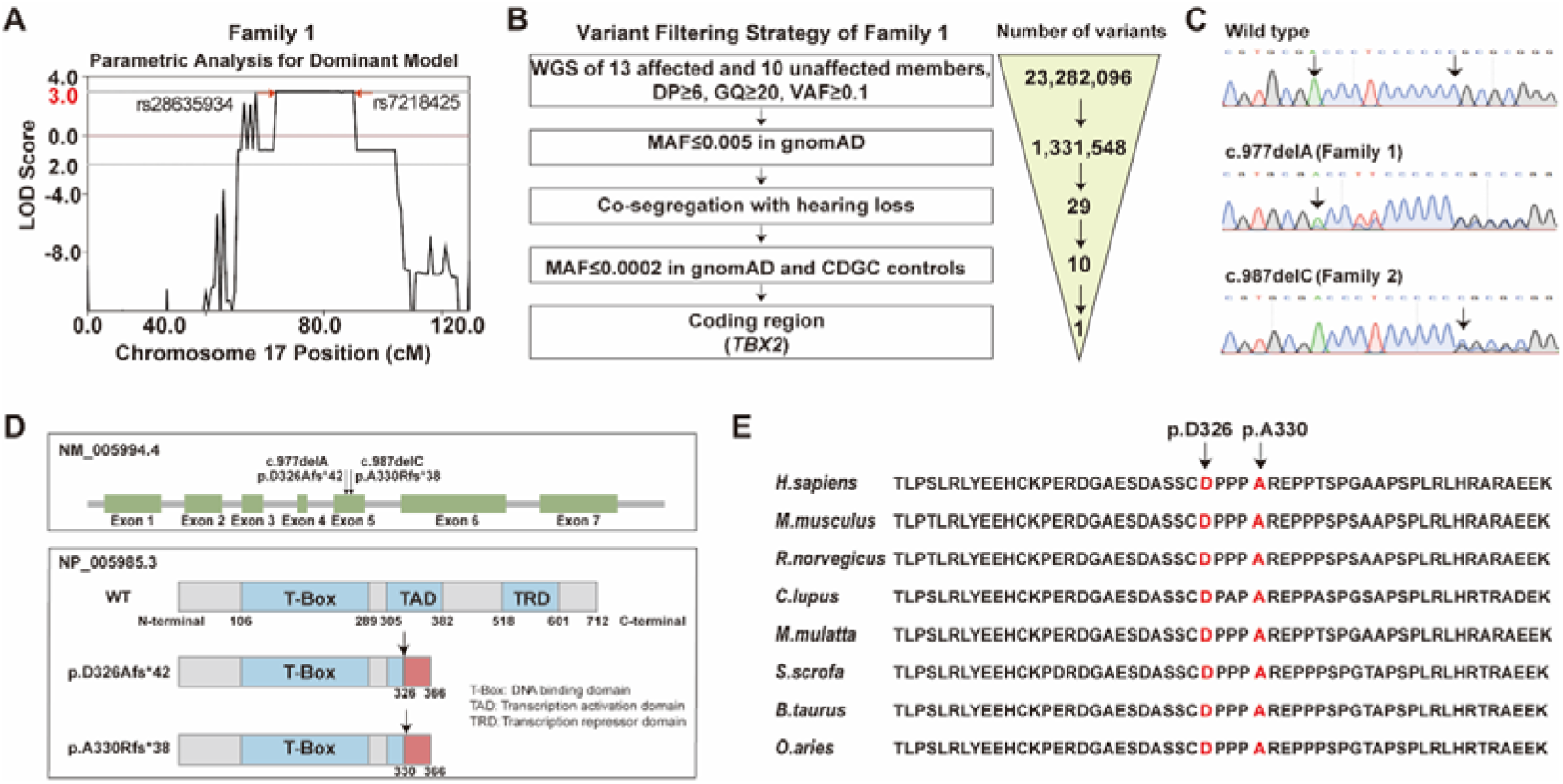
Identification of *TBX2* variants. A. A maximum LOD score of 3.01 on chromosome 17q23.2 in Family 1 by linkage analysis. B. Stepwise strategy for filtering WGS variants in Family 1. C. Sanger sequencing results of wild type and two *TBX2* variants in Family 1 and 2. D. Schematic diagram of the TBX2 protein highlighting positions of two variants marked by arrowheads, along with the truncated TBX2 protein sequence. E. Conservation analysis of amino acid sequences of two frameshift variants in different species.

Additionally, WGS performed on two patients and two unaffected members in Family 2 revealed another *TBX2* frameshift variant (NM_005994.4, c.987delC, p.Ala330Argfs*38) following the same filtering strategy (Table S2). This variant was co-segregation with the phenotype, confirmed via Sanger sequencing (Figure 2C). *TBX2* c.977delA was absent from the Genome Aggregation Database (gnomAD v4.0.0), while c.987delC was detected in one allele out of 1,415,242 alleles in the European (non-Finnish) population of gnomAD. Neither variant was detected in 7,395 ethnically matched CDGC controls.

Given that nystagmus was observed only in certain patients, linkage and WGS analysis were performed on Family 1 members with nystagmus as an independent phenotype. No potential candidate loci and genes were observed under the autosomal recessive model after filtering strategy (Table S3). Six shared variants were identified among the patients in autosomal dominant pattern (*PRKD3*, *DYNC2LI1*, *FAHD2A*, *OR5K3*, *TBX2*, *ZNF135*), with *TBX2* co-segregating within the extended family members (Table S4 and S5).

### Effect of *TBX2* variants in *in vitro* study

Two distinct *TBX2* frameshift variants (p.Asp326Alafs*42 and p.Ala330Argfs*38) were located in exon 5, leading to premature termination and the generation of new peptide segments sharing a common sequence of 33 amino acids (Figure S1). These variants were both within the transcription activation domain (Figure 2D), with the regions containing p.Asp326 and p.Ala330 highly conserved across species (Figure 2E). To evaluate the effects of these variants, wild type (WT) and mutant *TBX2* were transiently transfected in HEK293T and Hela cells. The destructive effects of these two variants on protein integrity were confirmed at the cellular level, with no significant change in protein expression (Figure 3A). The mutant did not alter TBX2 nuclear localization (Figure 3B), but exhibited significant reduced transcriptional activity compared to the WT protein (Figure 3C).

**Figure 3.**
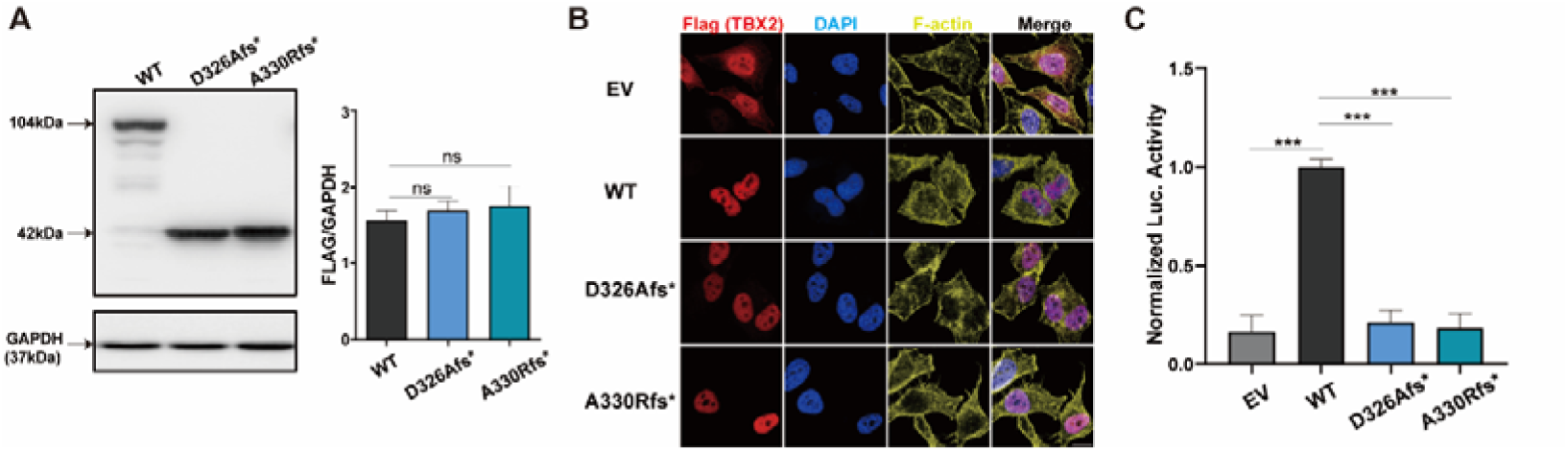
The effect of *TBX2* variants *in vitro*. A. The expression effect of p.Asp326Alafs*42 and p.Ala330Argfs*38 variants were confirmed by Western blot. B. Cellular localization of TBX2 proteins in Hela cells. C. The analysis of transcriptional activity of TBX2 variants using a dual-luciferase system. Scale bars, 15 μm. Data are presented as mean ± SEM. *** *p* < 0.001; ns: non-significant.

### Functional study of heterozygous *Tbx2* knockout mice

To assess the potential consequences of *TBX2* dysfunction, CRISPR/Cas9 was used to generate *Tbx2* knockout mice (*Tbx2^+/−^*) by targeting sgRNA-1 and −2 (Figure 4A). *Tbx2^-/-^* mice experienced early embryonic lethality due to cardiac insufficiency (19). *Tbx2^+/−^* mice exhibited normal hearing at postnatal day70 (P70), but showed progressively increasing in auditory thresholds with age. At P100, *Tbx2^+/−^* mice displayed significant auditory thresholds increases at 11.3 and 22.6 kHz compared to WT littermates, with further deterioration across multiple frequencies at P120 and P150 (Figure 4B). Prestin, an OHC marker, exhibited ectopic expression in IHCs of *Tbx2^+/−^* mice at P7, while it was specifically expressed in OHCs in WT mice (Figure 4C). The number of Prestin+ IHCs in *Tbx2^+/−^*mice gradually declined from P7 to P42, with significantly higher numbers at P7 compared to P42 (Figure 4D). Overall, heterozygous *Tbx2* ablation disrupted normal IHC development and resulted in late-onset, progressive hearing loss in mice, reflecting the clinical phenotype observed in humans.

**Figure 4.**
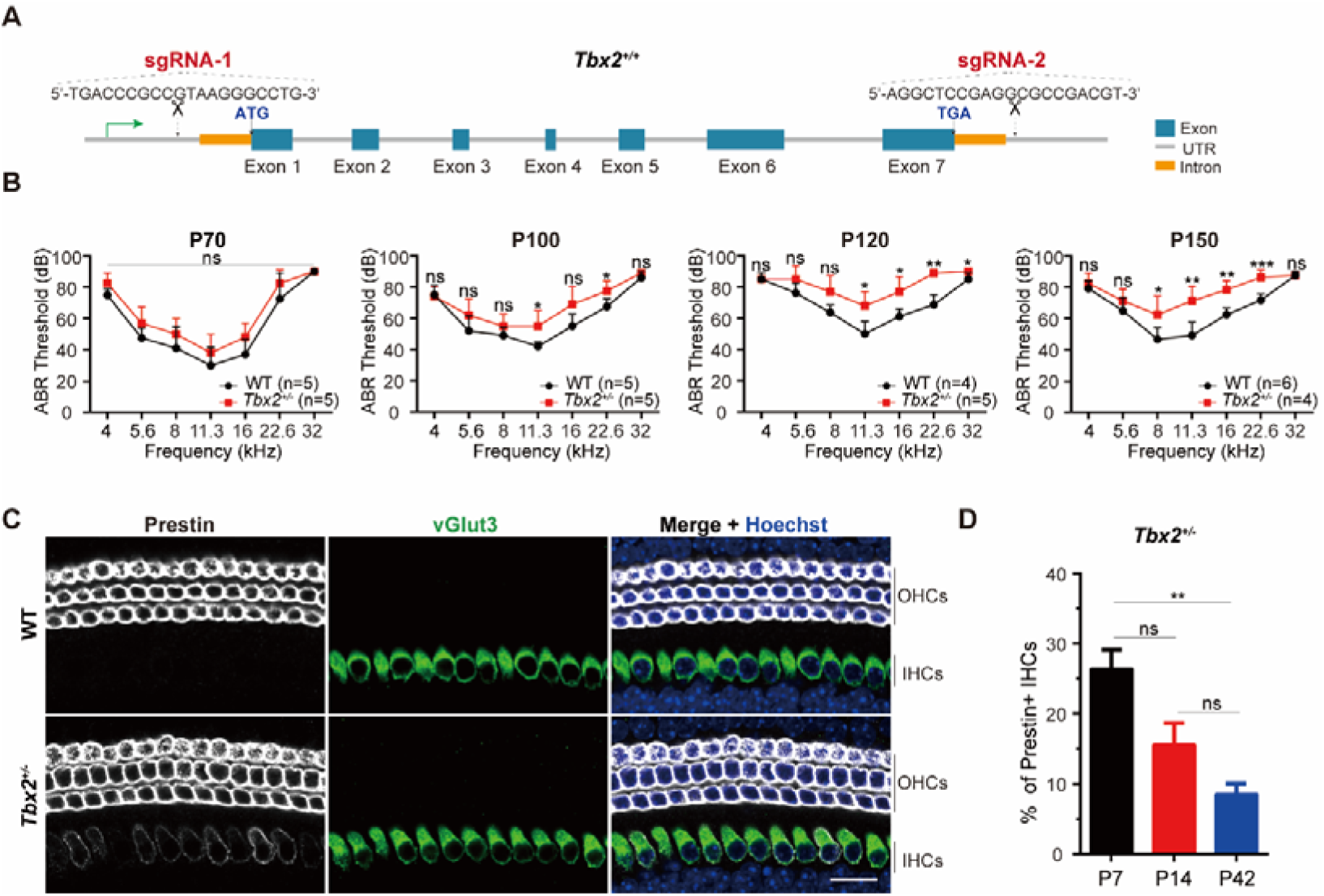
Functional study of heterozygous *Tbx2* knockout mice. A. Generation of germline *Tbx2^+/−^* mice by CRISPR/Cas9, involving the deletion of whole gene between sgRNA-1 and sgRNA-2 targeted sites. B. Auditory brainstem response (ABR) threshold comparison between wild type (WT) and *Tbx2^+/−^* mice from P70 to P150. C. Immunohistochemistry staining for Prestin (White) and vGlut3 (Green) in cochleae from WT and *Tbx2^+/−^* mice at P7 in apical turn. Scale bars, 20 μm. D. Quantification of Prestin+ IHC in *Tbx2^+/−^*mice at P7, P14, and P42. Data are presented as mean ± SEM. * *p* < 0.05; ** *p* < 0.01; *** *p* < 0.001; ns: non-significant.

## Discussion

In this study, we identified two frameshift variants of *TBX2* in two unrelated autosomal dominant Chinese families with late-onset progressive hearing loss, and nystagmus with incomplete penetrance. This finding underscores the significance of TBX2 in auditory function and its potential role in vestibular nystagmus.

The presence of nystagmus in only a subset of patients suggests two possibilities: environmental factors, which are less likely as they typically do not accompany hearing loss in two independent pedigrees, or that nystagmus and hearing loss are genetically distinct. Known causative genes for nystagmus were excluded, and *TBX2* was identified as the only candidate gene after co-segregation analysis in both families. Horizontal nystagmus, observed in partially subjects with hearing loss, is characteristic of both peripheral and central vestibular nystagmus. Given the lack of abnormalities detected in the central nervous system and visual system, we speculate that the nystagmus could be attributed to damage to the vestibular hair cells within the inner ear. Although TBX2 expression has been confirmed in vestibular tissues, its role in vestibular function remains to be elucidated (29, 30). Our findings provide a basis for investigating the important role of TBX2 in vestibular development.

We retrieved 306 syndromes with hearing loss and nystagmus as minor phenotypes from HPO, OMIM, Orphanet, and DECIPHER, but found no diseases with hearing loss and nystagmus as major phenotypes. Therefore, we propose that variants in *TBX2* are associated with a new autosomal-dominant disease with hearing loss and nystagmus with incomplete penetrance. Future genetic screenings of additional *TBX2* families will provide important clues about the incomplete penetrance of nystagmus, which may be influenced by genetic modification or allelic imbalance.

The *TBX2* gene encodes a transcription factor that belongs to the family of T-box factor proteins that bind to DNA. Several missense and stop-gain variants of *TBX2* have been associated with conotruncal heart defects, vertebral anomalies, variable endocrine, and osteochondrodysplasia (25–27). However, in this study, none of the affected members exhibited these phenotypes, suggesting that each variant type has distinct effects on the protein structure and function. The frameshifts identified in this study are located in the transcription activation domain, leading to the loss of the C-terminal peptide segment and potentially various clinical phenotypes.

In this study, the *TBX2* mutant did not alter protein expression and localization but resulted in reduced transcriptional activity, indicating a possible loss of function in *TBX2*. Our results show a correlation between *Tbx2* disruption in knockout mice and late-onset progressive hearing loss phenotype, suggesting that *TBX2* variants might be related to the haploinsufficiency. The dominant-negative effect cannot be excluded, as a novel peptide was generated in two *TBX2* frameshift variants. To establish a robust connection between the identified variants and the disease, it is crucial to employ knock-in mouse models and humanized knock-in models carrying these variants. These models will help elucidate the molecular mechanisms underlying the pathogenic effects of gene variants.

Tbx2 is closely related to the formation and development of the cochlea and considered a key regulatory factor for inner hair cell development and maturation (3–5). A decreasing trend in Prestin levels in IHCs from P7 to P42 upon *Tbx2* absence suggests its relevance to progressive hearing loss in mice and humans. While Tbx2 is a master regulator for IHCs development, its widespread expression throughout the inner ear implies additional functional roles beyond this domain. These roles may include essential functions in vestibular or broader auditory processes. Ectopic Prestin expression is maintained in the IHCs of conditional homozygous *Tbx2* mutants (12), suggesting that some aged IHCs may require less Tbx2 to maintain their adult fate.

In summary, our data indicate that heterozygous *TBX2* frameshift variants are the genetic cause of late-onset progressive hearing loss and incomplete penetrance of horizontal oscillatory nystagmus. Further investigation into other cases is warranted to characterize this novel disorder. Heterozygous *Tbx2* knockout mice showed progressive hearing loss, ectopic expression of Prestin in IHCs, and a gradual decrease in expression. Our findings will aid in elucidating the role of TBX2 in the auditory system and provide molecular diagnostics and genetic counseling for individuals at risk of or affected by this condition.

## Data Availability

All data are included in the main text and supplementary material appendix. In light of protecting patients’ privacy, please contact the author to request access to the information of Family 1 and 2.

## Acknowledgments

We would like to express our deepest gratitude to all the patients who participated in this study. We thank laboratory members for the helpful discussion.

## Author contributions

Conceptualization: all authors; Funding Acquisition: W.H., H.Y.; Investigation: W.H., Y.W., X.L., L.W., W.X., M.C., F.B., L.L., M.Z., Y.L.; Writing-original draft: W.H., J.C.; Writing-review and editing: all authors; Supervision: J.C., Z.L., H.Y.

## Ethics Declaration

Informed consent was obtained from all participants. This study was approved by the Institutional Review Board of West China Hospital.

