## Supplementary material for "Frameshift variants in *TBX2* underlie autosomal-dominant hearing loss with incomplete penetrance of nystagmus"

**Supplementary Results**

**Figure S1**

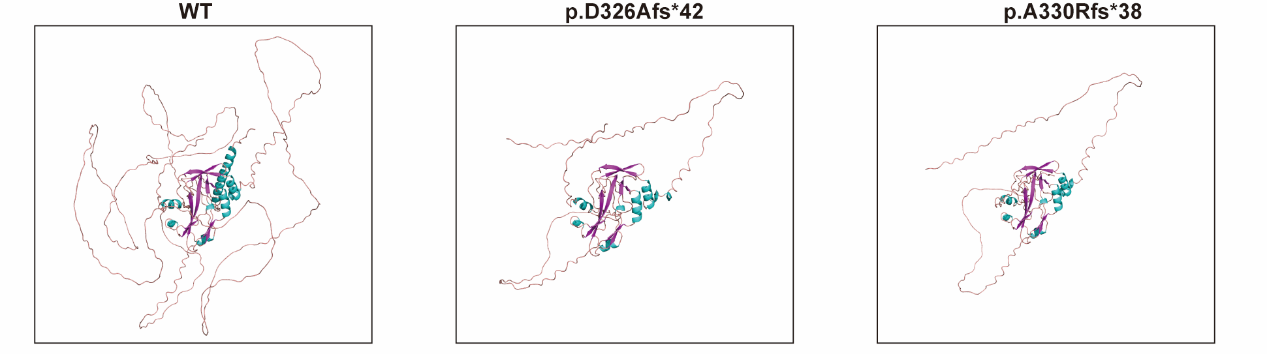

**Figure S1.** 3-D structure modeling of *TBX2* two variants (p.D326Afs*42, p.A330Rfs*38) identified in family 1 and family 2, respectively, based on the wild type structure predicted by AlphaFold. Figures were made with PyMOL.

**Table S1. Whole-genome sequencing and variant filtering strategy adapted to prioritize causative variant in Family 1.**

| Variant filtering process | | | Family 1 | | | | | | | | | | | | |
| --- | --- | --- | --- | --- | --- | --- | --- | --- | --- | --- | --- | --- | --- | --- | --- |
|  | Ⅲ:7 | | | Ⅳ:11 | Ⅳ:14 | Ⅳ:19 | Ⅳ:22 | Ⅳ:23 | Ⅴ:11 | Ⅴ:18 | Ⅴ:19 | Ⅴ:20 | Ⅴ:22 | Ⅴ:23 | Ⅴ:24 |
| DP≥6, GQ≥20, VAF≥0.1 | 5,092,120 | | | 5,020,554 | 5,194,718 | 5,118,127 | 5,222,963 | 5,023,559 | 5,178,040 | 5,258,876 | 5,262,085 | 5,144,839 | 5,269,993 | 5,262,125 | 5,211,392 |
| MAF≤0.005 in gnomAD | 276,531 | | | 273,474 | 275,416 | 268,258 | 291,667 | 260,963 | 276,970 | 275,734 | 284,943 | 272,731 | 286,579 | 285,657 | 277,352 |
| Coding variants, Nonsynonymous variants | 530 | | | 516 | 532 | 491 | 510 | 549 | 475 | 537 | 582 | 518 | 635 | 587 | 588 |
| Heterozygous variants | 371 | | | 360 | 348 | 353 | 385 | 391 | 374 | 394 | 380 | 392 | 398 | 402 | 348 |
| Sharing in all affected family members | | 2 | | | | | | | | | | | | | |
| Not detected in unaffected family members | | 1 (*TBX2*) | | | | | | | | | | | | | |

DP: Read Depth; GQ: Genotype Quality; VAF: Variant Allele Frequency; MAF: Minor Allele Frequency

**Table S2. Candidate variant detected in Family 2.**

| Gene | Genome position (GRCh38) | Nucleotide change | Amino acid change | AF_popmax  in gnomAD | SIFT | POLYPHEN-2 | Mutation  Taster |
| --- | --- | --- | --- | --- | --- | --- | --- |
| *ZFYVE9* | 1:52303877 | NM_004799.4:c.3390G>A | NP_004790.2:p.Met1130Ile | unknown | 0.041 | 0.94 | Damaging |
| *EFNA1* | 1:155131352 | NM_004428.3:c.106G>A | NP_004419.2:p.Asp36Asn | unknown | 0.035 | 0.998 | Damaging |
| *ACER3* | 11:76926601 | NM_018367.7:c.148G>A | NP_060837.3:p.Gly50Ser | unknown | 0.093 | 1 | Damaging |
| *ACAT1* | 11:108131937 | NM_000019.4:c.103T>C | NP_000010.1:p.Ser35Pro | unknown | 0.026 | 0.004 | Damaging |
| *FOXRED1* | 11:126276075 | NM_017547.4:c.827G>A | NP_060017.1:p.Ser276Asn | unknown | 0.156 | 0.93 | Damaging |
| *ENO2* | 12:6917581 | NM_001975.3:c.311C>G | NP_001966.1:p.Ser104Cys | unknown | 0.001 | 0.95 | Damaging |
| *FLT3* | 13:28052585 | NM_004119.3:c.574C>G | NP_004110.2:p.Pro192Ala | 0.0000147 | 0 | 1 | Tolerated |
| *NUP50* | 22:45171604 | NM_007172.4:c.74G>A | NP_009103.2:p.Gly25Glu | unknown | 0.002 | 0.998 | Damaging |
| *ATG4C* | 1:62816716 | NM_032852.4:c.302C>A | NP_116241.2:p.Ser101Ter | unknown | NA | NA | NA |
| *TBX2* | 17:61404700 | NM_005994.4:c.987delC | NP_005985.3:p.Ala330Argfs*38 | unknown | NA | NA | NA |

SIFT: Sorts Intolerant From Tolerant, Deleterious < 0.05, Tolerated ≥ 0.05; POLYPHEN-2: Polymorphism Phenotyping v2, Possibly damaging > 0.909, 0.447 ≤ Possibly damaging < 0.909, Benign ≤ 0.446.

NA: not available.

**Table S3. Screening for pathogenic genes in patients carrying nystagmus in Family 1 with autosomal recessive inheritance pattern.**

| Variant filtering process | Family 1 | | | | |
| --- | --- | --- | --- | --- | --- |
|  | Ⅳ:22 | Ⅳ:23 | Ⅴ:20 | Ⅴ:21 | Ⅴ:22 |
| DP≥6, GQ≥20, VAF≥0.1 | 5,222,963 | 5,023,559 | 5,144,839 | 5,157,985 | 5,269,993 |
| MAF≤0.005 in gnomAD | 291,667 | 260,963 | 272,731 | 256,432 | 286,579 |
| Coding variants, Nonsynonymous variants | 510 | 549 | 518 | 634 | 635 |
| Homozygous or compound heterozygote variants shared in all affected family members | 9 (2 genes) | | | | |
| Not present in unaffected family members | 0 | | | | |

DP: Read Depth; GQ: Genotype Quality; VAF: Variant Allele Frequency; MAF: Minor Allele Frequency

**Table S4. Screening for pathogenic genes in patients carrying nystagmus in Family 1 with autosomal dominant inheritance pattern.**

| Variant filtering process | Family 1 | | | | |
| --- | --- | --- | --- | --- | --- |
| WGS | Ⅳ:22 | Ⅳ:23 | Ⅴ:20 | Ⅴ:21 | Ⅴ:22 |
| DP≥6, GQ≥20, VAF≥0.1 | 5,222,963 | 5,023,559 | 5,144,839 | 5,157,985 | 5,269,993 |
| MAF≤0.005 in gnomAD | 291,667 | 260,963 | 272,731 | 256,432 | 286,579 |
| Coding variants, Nonsynonymous variants | 510 | 549 | 518 | 634 | 635 |
| Heterozygous variants | 385 | 391 | 392 | 458 | 398 |
| Present in all affected family members | 6 | | | | |
| Segregation analysis, gene/disease association | 1 (*TBX2*) | | | | |

DP: Read Depth; GQ: Genotype Quality; VAF: Variant Allele Frequency; MAF: Minor Allele Frequency

**Table S5. 6 candidate variants shared in patients with nystagmus in Family 1.**

| Gene | Genome position (GRCh38) | Nucleotide change | Amino acid change | AF_popmax in gnomAD | SIFT | POLYPHEN-2 | MutationTaster |
| --- | --- | --- | --- | --- | --- | --- | --- |
| *PRKD3* | 2:37279872 | NM_005813.6:c.1046T>G | NP_005804.1:p.Ile349Arg | 0.0013503 | 0.374 | 0.006 | Damaging |
| *DYNC2LI1* | 2:43794617 | NM_016008.4:c.481T>G | NP_057092.2:p.Trp161Gly | 0.0001923 | 0.33 | 0.118 | Damaging |
| *FAHD2A* | 2:95410967 | NM_016044.3:c.626A>G | NP_057128.2:p.Lys209Arg | 0.0007698 | 0 | 1 | Damaging |
| *OR5K3* | 3:98391443 | NM_001005516.1:c.778C>G | NP_001005516.1:p.Pro260Ala | unknown | 0.001 | 0.998 | Damaging |
| *TBX2* | 17: 61404695 | NM_005994.4:c.977delA | NP_005985.3:p.Asp326Alafs*42 | unknown | NA | NA | NA |
| *ZNF135* | 19: 58067388 | NM_001289401.2:c.904G>A | NP_001276330.1:p.Glu302Lys | 0.001934 | 0.006 | 0.999 | Tolerated |

SIFT: Sorts Intolerant From Tolerant, Deleterious < 0.05, Tolerated ≥ 0.05; POLYPHEN-2: Polymorphism Phenotyping v2, Possibly damaging > 0.909, 0.447 ≤ Possibly damaging < 0.909, Benign ≤ 0.446.

NA: not available.

**Video S1. Nystagmus video of Ⅴ:22 in family 1, recorded in 2022.**
